# Estimating COVID-19 Prevalence in the United States: A Sample Selection Model Approach

**DOI:** 10.1101/2020.04.20.20072942

**Authors:** David Benatia, Raphael Godefroy, Joshua Lewis

## Abstract

**Background:** Public health efforts to determine population infection rates from coronavirus disease 2019 (COVID-19) have been hampered by limitations in testing capabilities and the large shares of mild and asymptomatic cases. We developed a methodology that corrects observed positive test rates for non-random sampling to estimate population infection rates across U.S. states from March 31 to April 7.

**Methods:** We adapted a sample selection model that corrects for non-random testing to estimate population infection rates. The methodology compares how the observed positive case rate vary with changes in the size of the tested population, and applies this gradient to infer total population infection rates. Model identification requires that variation in testing rates be uncorrelated with changes in underlying disease prevalence. To this end, we relied on data on day-to-day changes in completed tests across U.S. states for the period March 31 to April 7, which were primarily influenced by immediate supply-side constraints. We used this methodology to construct predicted infection rates across each state over the sample period. We also assessed the sensitivity of the results to controls for state-specific daily trends in infection rates.

**Results:** The median population infection rate over the period March 31 to April 7 was 0.9% (IQR 0.64 1.77). The three states with the highest prevalence over the sample period were New York (8.5%), New Jersey (7.6%), and Louisiana (6.7%). Estimates from mod-els that control for state-specific daily trends in infection rates were virtually identical to the baseline findings. The estimates imply a nationwide average of 12 population infections per diagnosed case. We found a negative bivariate relationship (corr. = -0.51) between total per capita state testing and the ratio of population infections per diagnosed case.

**Interpretation:** The effectiveness of the public health response to the coronavirus pandemic will depend on timely information on infection rates across different regions. With increasingly available high frequency data on COVID-19 testing, our methodology could be used to estimate population infection rates for a range of countries and subnational districts. In the United States, we found widespread undiagnosed COVID-19 infection. Expansion of rapid diagnostic and serological testing will be critical in preventing recurrent unobserved community transmission and identifying the large numbers individuals who may have some level of viral immunity.

**Funding:** Social Sciences and Humanities Research Council.

## 1 Introduction

In December 2019, several clusters of pneumonia cases were reported in the Chinese city of Wuhan. By early January, Chinese scientists had isolated a novel coronavirus (SARS-CoV-2), later named coronavirus disease 2019 (COVID-19), for which a laboratory test was quickly developed. Despite efforts at containment through travel restrictions, the virus spread rapidly beyond mainland China. By April 7, more than 1.4 million cases had been reported in 182 countries and regions.

Our understanding of the progression and severity of the outbreak has been limited by constraints on testing capabilities. In most countries, testing has been limited to a small fraction of the population. As a result, the number of confirmed positive cases may grossly understate the population infection rate, given the large numbers of mild and asymptomatic cases that may go untested [1–5]. Moreover, testing has often been targeted to specific subgroups, such as individuals who were symptomatic or who were previously exposed to the virus, whose infection probability differs from that in the overall population [6, 7].^1^ Given this *sample selection bias*, it is impossible to infer overall disease prevalence from the share of positive cases among the tested individuals.

A further challenge to our understanding of the spread of outbreak has been the wide variation in per capita testing across jurisdictions due to different protocols and testing capabilities. For example, as of April 7, South Korea had conducted three times more tests than the United States on a per capita basis [8,9]. Large differences in testing rates also exist at the subnational level. For example, per capita testing in the state of New York was nearly two times higher than in neighboring New Jersey [8]. Because the severity of sample selection bias depends on the extent of testing, these disparities create large uncertainty regarding the relative disease prevalence across jurisdictions, and may contribute to the wide differences in estimated case fatality rates [10, 11].

In this study, we implemented a procedure that corrects observed infection rates among tested individuals for non-random sampling to calculate population disease prevalence. A large body of empirical work in economics has been devoted to the problem of sample selection and researchers have developed estimation procedures to correct for non-random sampling [12–17]. Our methodology builds on these insights to correct observed infection rates for non-random selection into COVID-19 testing.

Our procedure compares how the observed infection rate varied as a larger share of the population was tested, and uses this gradient to infer disease prevalence in the overall population. Because investments in testing capacity may respond endogenously to local disease conditions, however, model identification requires that we find a source of variation in testing rates COVID-19 that is unrelated to the underlying population prevalence. To this end, we relied on high frequency day-to-day changes in completed tests across U.S. states, which were primarily driven by immediate supply-side limitations rather than the more gradual evolution of local disease prevalence. We used this procedure to correct for selection bias in observed infection rates to calculate population disease prevalence across U.S. states from March 31 to April 7.

## 2 Methodology

### 2.1 Theory

To evaluate population disease prevalence, we developed a simple selection model for COVID-19 testing and used the framework to link observed rates of positive tests to population disease prevalence. We considered a stable population, denoting *A* and *B* as the numbers of sick and healthy individuals, respectively. Let *p*_*n*_ denote the probability that a sick person is tested and *q*_*n*_ the probability that a healthy person is tested, given a total number of tests, *n*. Thus, we have:

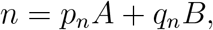

and the number of positive tests is:

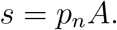

This simple framework highlights how non-random testing will bias estimates of the population disease prevalence. Using Bayes’ rule, we can write the relative probability of testing as the following:

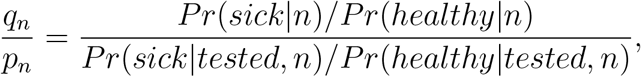

which is equal to one if tests are randomly allocated, *Pr*(*sick*|*tested, n*) = *Pr*(*sick*|*n*). When testing is targeted to individuals who are more likely to be sick, we have *Pr*(*sick*|*tested, n*) *> Pr*(*sick*|*n*) and *Pr*(*healthy*|*tested, n*) *< Pr*(*healthy*|*n*), so the ratio will fall between zero and one. In this scenario, the ratio of sick to healthy people in the sample, *p*_*n*_*A/q*_*n*_*B*, will exceed the ratio in the overall population, *A/B*.

We specified the following functional form for the relative probability of testing:

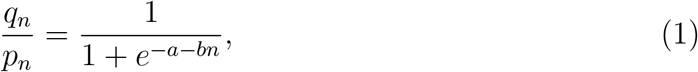

which is in [0,1] for *−a − bn ≤* 0. The term *e*^*−a−bn*^ *>* 0 reflects the fact that testing has been targeted towards higher risk populations, with the intercept, *−a*, capturing the severity of selection bias when testing is limited. Meanwhile, the coefficient *b >* 0 identifies how selection bias decreases with *n* as the ratio *q*_*n*_*/p*_*n*_ approaches one. Intuitively, as testing expands, the sample will become more representative of the overall population, and the selection bias will diminish.

Combining both equations, we have:

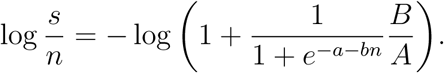

We used the fact that the ratio of negative to positive tests is much larger than one to make the following approximation:^2^

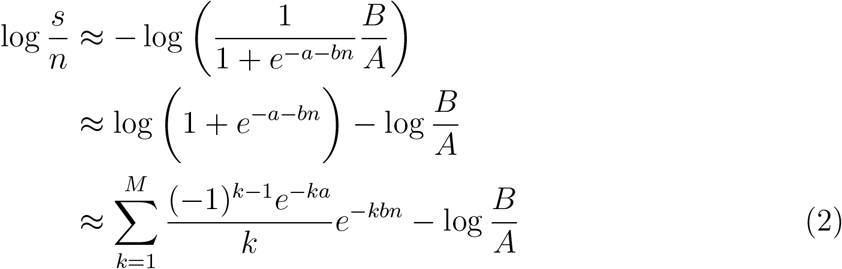

Given a change in the number of tests conducted in a particular population, *n*_1_ to *n*_2_, equation (2) implies the following change in the share of positive tests:

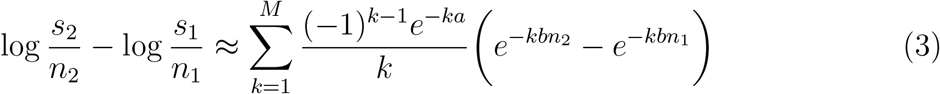

### 2.2 Model Estimation and Identification

Our empirical model was derived from equation (3). We used information on testing across states *i* on day *t* to estimate the following equation:

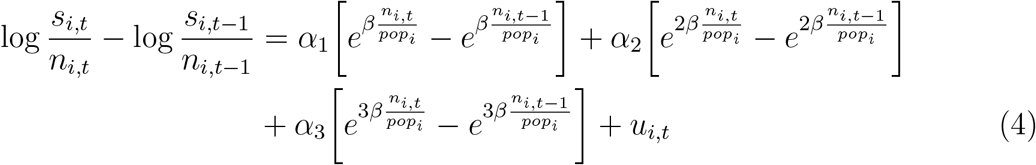

where *n*_*i,t*_ is the number of tests on day *t, s*_*i,t*_ is the share of positive tests, and *pop*_*i*_ is the state population. The term *u*_*i,t*_ is an error which we assumed to follow a Gaussian distribution with mean zero and unknown variance. We restricted the model to a cubic approximation of the function in equation (4), since higher order terms were found to be statistically insignificant. This approximation is supported by graphical evidence depicted below. We estimated equation (4) by maximum likelihood.

For model identification, we required that day-to-day changes in the number of tests be uncorrelated with the error term, *u*_*i,t*_. In practice, this assumption implies that daily changes in underlying population disease prevalence cannot be systematically related to day-to-day changes in testing. Our identification assumption is supported by at least three pieces of evidence. First, severe constraints on state testing capacity have caused a significant backlog in cases, so that changes in the number of daily tests primarily reflects changes in local capacity rather than changes in demand for testing. Second, because our analysis focuses on high frequency day-to-day changes in outcomes, there is limited scope for large evolution in underlying disease prevalence. Finally, in robustness exercises, we augmented the basic model to include state fixed effects, thereby allowing for state-specific exponential growth in underlying disease prevalence from one day to the next. These additional controls did not alter the main empirical findings.

To recover estimates of population infection rates, 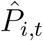, in state *i* at date *t*, we combined the estimates from equation (4) and set *n* = *pop*_*i*_ according to the following equation:

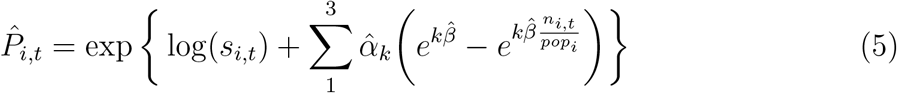

We then used the Delta-method to estimate the confidence interval for 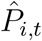.

### 2.3 Data

The analysis was based on daily information on total tests results (positive plus negative) and total positive test results across U.S. states for the period March 31 to April 7. These data were obtained from the COVID Tracking Project, a site that was launched by journalists from The Atlantic to publish high-quality data on the outbreak in the United Stated [8]. The data were originally compiled primarily from state public health authorities, occasionally supplemented by information from news reporting, official press conferences, or message from officials released on facebook or twitter. We focused on the recent period to limit errors associated with previous changes in state reporting practices. We supplemented this information with data on total state population from the census [18].

## 3 Results

Table 1 (Model 1) reports the estimated coefficients from equation (4). Model 2 and Model 3 present the estimation results from models with state fixed effects (Model 2), and for the subsample of state-day observations with a positive test ratio smaller than 0.5 (Model 3).

**Table 1:**
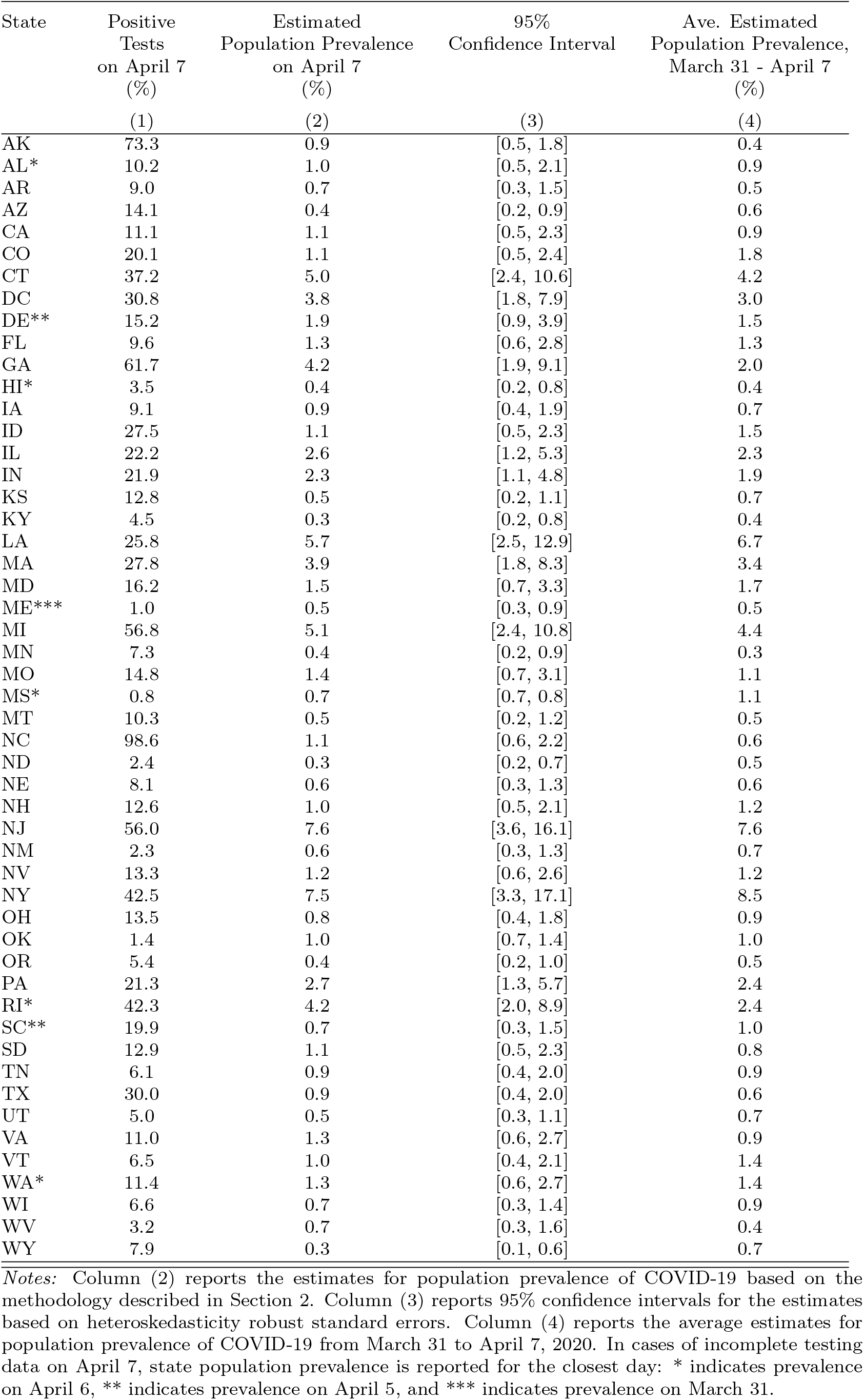
Estimated Population Infection Rates for COVID-19

| State | Positive<br>Tests<br>on April 7<br>(%) | Estimated<br>Population Prevalence<br>on April 7<br>(%) | 95%<br>Confidence Interval | Ave. Estimated<br>Population Prevalence,<br>March 31 - April 7<br>(%) |
| --- | --- | --- | --- | --- |
|  | (1) | (2) | (3) | (4) |
| AK | 73.3 | 0.9 | [0.5, 1.8] | 0.4 |
| AL* | 10.2 | 1.0 | [0.5, 2.1] | 0.9 |
| AR | 9.0 | 0.7 | [0.3, 1.5] | 0.5 |
| AZ | 14.1 | 0.4 | [0.2, 0.9] | 0.6 |
| CA | 11.1 | 1.1 | [0.5, 2.3] | 0.9 |
| CO | 20.1 | 1.1 | [0.5, 2.4] | 1.8 |
| CT | 37.2 | 5.0 | [2.4, 10.6] | 4.2 |
| DC | 30.8 | 3.8 | [1.8, 7.9] | 3.0 |
| DE** | 15.2 | 1.9 | [0.9, 3.9] | 1.5 |
| FL | 9.6 | 1.3 | [0.6, 2.8] | 1.3 |
| GA | 61.7 | 4.2 | [1.9, 9.1] | 2.0 |
| HI* | 3.5 | 0.4 | [0.2, 0.8] | 0.4 |
| IA | 9.1 | 0.9 | [0.4, 1.9] | 0.7 |
| ID | 27.5 | 1.1 | [0.5, 2.3] | 1.5 |
| IL | 22.2 | 2.6 | [1.2, 5.3] | 2.3 |
| IN | 21.9 | 2.3 | [1.1, 4.8] | 1.9 |
| KS | 12.8 | 0.5 | [0.2, 1.1] | 0.7 |
| KY | 4.5 | 0.3 | [0.2, 0.8] | 0.4 |
| LA | 25.8 | 5.7 | [2.5, 12.9] | 6.7 |
| MA | 27.8 | 3.9 | [1.8, 8.3] | 3.4 |
| MD | 16.2 | 1.5 | [0.7, 3.3] | 1.7 |
| ME*** | 1.0 | 0.5 | [0.3, 0.9] | 0.5 |
| MI | 56.8 | 5.1 | [2.4, 10.8] | 4.4 |
| MN | 7.3 | 0.4 | [0.2, 0.9] | 0.3 |
| MO | 14.8 | 1.4 | [0.7, 3.1] | 1.1 |
| MS* | 0.8 | 0.7 | [0.7, 0.8] | 1.1 |
| MT | 10.3 | 0.5 | [0.2, 1.2] | 0.5 |
| NC | 98.6 | 1.1 | [0.6, 2.2] | 0.6 |
| ND | 2.4 | 0.3 | [0.2, 0.7] | 0.5 |
| NE | 8.1 | 0.6 | [0.3, 1.3] | 0.6 |
| NH | 12.6 | 1.0 | [0.5, 2.1] | 1.2 |
| NJ | 56.0 | 7.6 | [3.6, 16.1] | 7.6 |
| NM | 2.3 | 0.6 | [0.3, 1.3] | 0.7 |
| NV | 13.3 | 1.2 | [0.6, 2.6] | 1.2 |
| NY | 42.5 | 7.5 | [3.3, 17.1] | 8.5 |
| OH | 13.5 | 0.8 | [0.4, 1.8] | 0.9 |
| OK | 1.4 | 1.0 | [0.7, 1.4] | 1.0 |
| OR | 5.4 | 0.4 | [0.2, 1.0] | 0.5 |
| PA | 21.3 | 2.7 | [1.3, 5.7] | 2.4 |
| RI* | 42.3 | 4.2 | [2.0, 8.9] | 2.4 |
| SC** | 19.9 | 0.7 | [0.3, 1.5] | 1.0 |
| SD | 12.9 | 1.1 | [0.5, 2.3] | 0.8 |
| TN | 6.1 | 0.9 | [0.4, 2.0] | 0.9 |
| TX | 30.0 | 0.9 | [0.4, 2.0] | 0.6 |
| UT | 5.0 | 0.5 | [0.3, 1.1] | 0.7 |
| VA | 11.0 | 1.3 | [0.6, 2.7] | 0.9 |
| VT | 6.5 | 1.0 | [0.4, 2.1] | 1.4 |
| WA* | 11.4 | 1.3 | [0.6, 2.7] | 1.4 |
| WI | 6.6 | 0.7 | [0.3, 1.4] | 0.9 |
| WV | 3.2 | 0.7 | [0.3, 1.6] | 0.4 |
| WY | 7.9 | 0.3 | [0.1, 0.6] | 0.7 |

*Notes:* Column (2) reports the estimates for population prevalence of COVID-19 based on the methodology described in Section 2. Column (3) reports 95% confidence intervals for the estimates based on heteroskedasticity robust standard errors. Column (4) reports the average estimates for population prevalence of COVID-19 from March 31 to April 7, 2020. In cases of incomplete testing data on April 7, state population prevalence is reported for the closest day: \* indicates prevalence on April 6, \*\* indicates prevalence on April 5, and \*\*\* indicates prevalence on March 31.

Figure 1 depicts the relationship between daily changes in the positive test rate and per capita testing, based on the relationship implied by equation (4), estimated across states for the period March 31 to April 7. Because 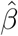 is *negative*, the upward sloping pattern implies a negative relationship between daily changes in testing and the share of positive tests. A symptom of selection bias is that variables that have no structural relationship with the dependent variable may appear to be significant [13]. Thus, these patterns strongly suggest non-random testing, since daily changes in testing should be unrelated to population disease prevalence except through a selection channel.^3^

**Figure 1:**
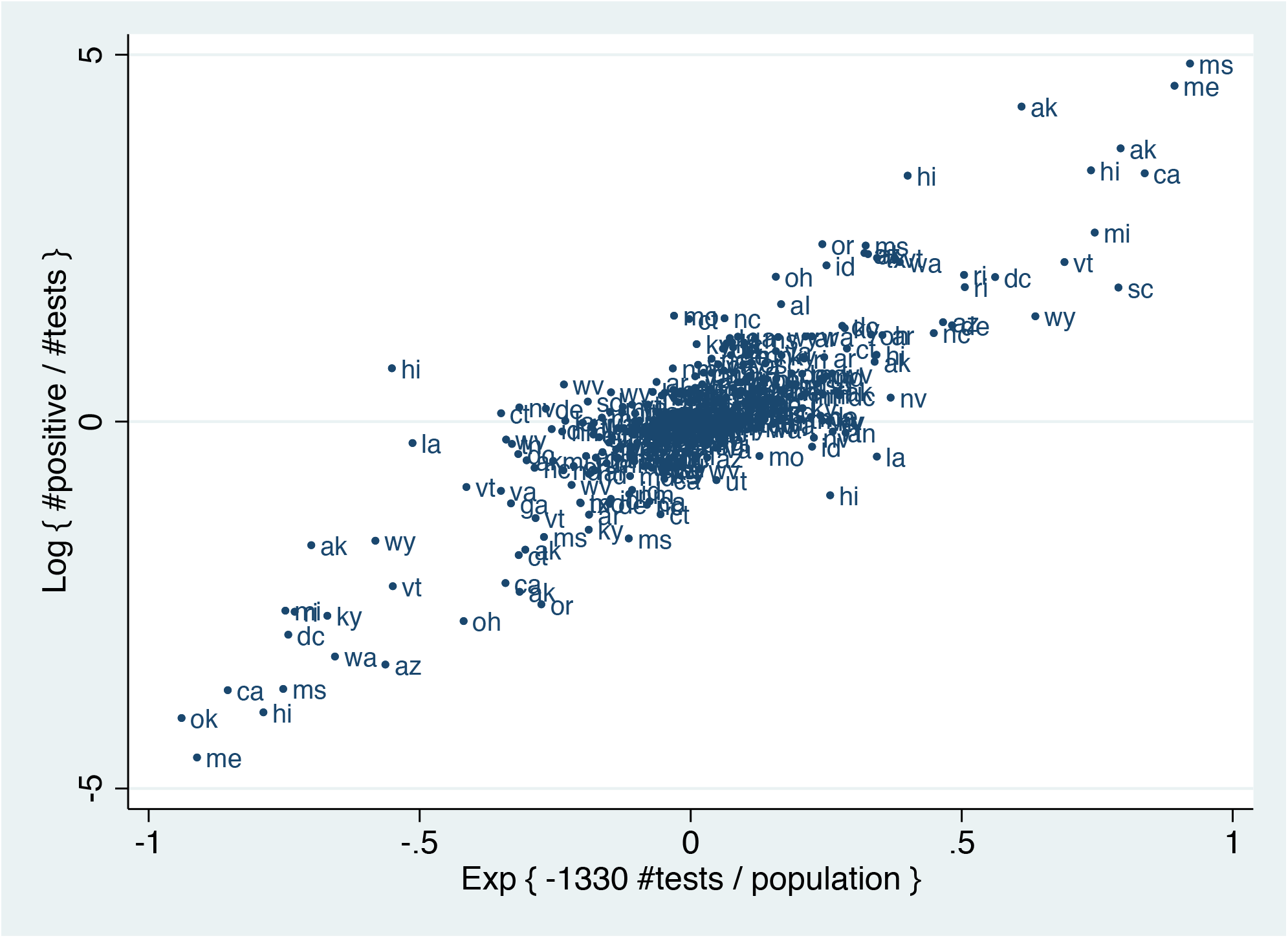
Daily Changes in Testing and the Share of Positive Cases. *Notes:* This figure reports the relationship between daily changes in the exponential of per capita testing and daily changes in the log share of positive tests, using the coefficient of *β* derived from the main estimates of equation (4).

Table 2 reports the results that adjust observed COVID-19 case rates for non-random testing based on the procedure described in Section 2. For reference, column (1) reports the observed positive test rate on April 7, 2020. Columns (2) and (3) report the adjusted rates for April 7 along with 95 percent confidence interval. The results suggest widespread undiagnosed cases of COVID-19. Estimated population prevalence ranged from 0.3 percent in Wyoming to 7.6 percent in New Jersey. To put these estimates in perspective, in New York state, which had conducted the most extensive testing in the nation, 0.7 percent of the population had tested positive for COVID-19 by April 7. Our estimates imply that 34 states had population case rates that exceeded the observed prevalence in New York.

**Table 2:** Robustness Exercises

|  | Estimated COVID-19 Prevalence<br>on April 7 |  |  |  |  |  | Estimated COVID-19 Prevalence<br>Average: March 31 - April 7 |  |  |
| --- | --- | --- | --- | --- | --- | --- | --- | --- | --- |
|  | Baseline<br>estimates |  | Add state<br>fixed effects |  | Restrict to<br>days w. < 50%<br>positive cases |  | Baseline<br>estimates | Add state<br>fixed<br>effects | Restrict to<br>days w. <<br>50% pos.<br>cases |
|  | Estimate | 95% CI | Estimate | 95% CI | Estimate | 95% CI |  |  |  |
|  | (1) | (2) | (3) | (4) | (5) | (6) | (7) | (8) | (9) |
| AK | 0.9 | [0.5, 1.8] | 1.0 | [0.6, 1.6] |  |  | 0.4 | 0.5 | 0.3 |
| AL* | 1.0 | [0.5, 2.1] | 1.0 | [0.6, 1.8] | 0.8 | [0.4, 1.9] | 0.9 | 0.9 | 0.8 |
| AR | 0.7 | [0.3, 1.5] | 0.7 | [0.4, 1.3] | 0.6 | [0.3, 1.3] | 0.5 | 0.6 | 0.5 |
| AZ | 0.4 | [0.2, 0.9] | 0.5 | [0.3, 0.8] | 0.4 | [0.2, 0.9] | 0.6 | 0.6 | 0.5 |
| CA | 1.1 | [0.5, 2.3] | 1.1 | [0.7, 2.0] | 0.9 | [0.4, 2.1] | 0.9 | 0.9 | 0.8 |
| CO | 1.1 | [0.5, 2.4] | 1.2 | [0.7, 2.1] | 0.9 | [0.4, 2.2] | 1.8 | 1.9 | 1.5 |
| CT | 5.0 | [2.4, 10.6] | 5.2 | [3.0, 9.1] | 4.2 | [1.8, 9.5] | 4.2 | 4.3 | 3.1 |
| DC | 3.8 | [1.8, 7.9] | 3.9 | [2.3, 6.7] | 3.2 | [1.4, 7.0] | 3.0 | 3.1 | 2.4 |
| DE** | 1.9 | [0.9, 3.9] | 2.0 | [1.1, 3.4] | 1.6 | [0.7, 3.5] | 1.5 | 1.6 | 1.3 |
| FL | 1.3 | [0.6, 2.8] | 1.4 | [0.8, 2.4] | 1.1 | [0.5, 2.5] | 1.3 | 1.3 | 1.1 |
| GA | 4.2 | [1.9, 9.1] | 4.3 | [2.5, 7.7] |  |  | 2.0 | 2.1 | 1.4 |
| HI* | 0.4 | [0.2, 0.8] | 0.4 | [0.2, 0.7] | 0.3 | [0.1, 0.7] | 0.4 | 0.5 | 0.4 |
| IA | 0.9 | [0.4, 1.9] | 0.9 | [0.5, 1.6] | 0.8 | [0.3, 1.7] | 0.7 | 0.7 | 0.6 |
| ID | 1.1 | [0.5, 2.3] | 1.1 | [0.6, 2.0] | 0.9 | [0.4, 2.1] | 1.5 | 1.6 | 1.3 |
| IL | 2.6 | [1.2, 5.3] | 2.7 | [1.6, 4.6] | 2.1 | [1.0, 4.8] | 2.3 | 2.4 | 1.9 |
| IN | 2.3 | [1.1, 4.8] | 2.4 | [1.4, 4.1] | 1.9 | [0.8, 4.3] | 1.9 | 2.0 | 1.6 |
| KS | 0.5 | [0.2, 1.1] | 0.5 | [0.3, 1.0] | 0.4 | [0.2, 1.0] | 0.7 | 0.7 | 0.6 |
| KY | 0.3 | [0.2, 0.8] | 0.4 | [0.2, 0.6] | 0.3 | [0.1, 0.7] | 0.4 | 0.4 | 0.3 |
| LA | 5.7 | [2.5, 12.9] | 5.9 | [3.2, 10.8] | 4.6 | [1.9, 11.3] | 6.7 | 6.9 | 5.0 |
| MA | 3.9 | [1.8, 8.3] | 4.0 | [2.3, 7.1] | 3.2 | [1.4, 7.3] | 3.4 | 3.6 | 2.8 |
| MD | 1.5 | [0.7, 3.3] | 1.6 | [0.9, 2.8] | 1.3 | [0.6, 2.9] | 1.7 | 1.8 | 1.4 |
| ME*** | 0.5 | [0.3, 0.9] | 0.5 | [0.4, 0.8] | 0.5 | [0.2, 0.9] | 0.5 | 0.5 | 0.5 |
| MI | 5.1 | [2.4, 10.8] | 5.3 | [3.0, 9.2] |  |  | 4.4 | 4.6 | 3.2 |
| MN | 0.4 | [0.2, 0.9] | 0.4 | [0.3, 0.8] | 0.4 | [0.2, 0.8] | 0.3 | 0.3 | 0.3 |
| MO | 1.4 | [0.7, 3.1] | 1.5 | [0.9, 2.6] | 1.2 | [0.5, 2.7] | 1.1 | 1.2 | 0.9 |
| MS* | 0.7 | [0.7, 0.8] | 0.7 | [0.7, 0.8] | 0.7 | [0.7, 0.8] | 1.1 | 1.1 | 0.9 |
| MT | 0.5 | [0.2, 1.2] | 0.6 | [0.3, 1.0] | 0.5 | [0.2, 1.1] | 0.5 | 0.5 | 0.4 |
| NC | 1.1 | [0.6, 2.2] | 1.2 | 0.7, 1.9 |  |  | 0.6 | 0.7 | 0.5 |
| ND | 0.3 | [0.2, 0.7] | 0.3 | [0.2, 0.6] | 0.3 | [0.1, 0.6] | 0.5 | 0.5 | 0.4 |
| NE | 0.6 | [0.3, 1.3] | 0.6 | [0.3, 1.1] | 0.5 | [0.2, 1.1] | 0.6 | 0.6 | 0.5 |
| NH | 1.0 | [0.5, 2.1] | 1.0 | [0.6, 1.8] | 0.8 | [0.4, 1.9] | 1.2 | 1.3 | 1.0 |
| NJ | 7.6 | [3.6, 16.1] | 7.9 | [4.5, 13.8] |  |  | 7.6 | 7.9 | 6.0 |
| NM | 0.6 | [0.3, 1.3] | 0.6 | [0.3, 1.1] | 0.5 | [0.2, 1.1] | 0.7 | 0.7 | 0.5 |
| NV | 1.2 | [0.6, 2.6] | 1.3 | [0.7, 2.2] | 1.0 | [0.5, 2.4] | 1.2 | 1.2 | 1.0 |
| NY | 7.5 | [3.3, 17.1] | 7.9 | [4.3, 14.4] | 6.2 | [2.6, 14.9] | 8.5 | 8.8 | 7.0 |
| OH | 0.8 | [0.4, 1.8] | 0.9 | [0.5, 1.5] | 0.7 | [0.3, 1.6] | 0.9 | 0.9 | 0.7 |
| OK | 1.0 | [0.7, 1.4] | 1.0 | [0.8, 1.3] | 0.9 | [0.6, 1.4] | 1.0 | 1.0 | 0.9 |
| OR | 0.4 | [0.2, 1.0] | 0.5 | [0.3, 0.8] | 0.4 | [0.2, 0.9] | 0.5 | 0.5 | 0.4 |
| PA | 2.7 | [1.3, 5.7] | 2.8 | [1.6, 4.9] | 2.3 | [1.0, 5.1] | 2.4 | 2.5 | 2.0 |
| RI* | 4.2 | [2.0, 8.9] | 4.4 | [2.5, 7.6] | 3.5 | [1.6, 8.0] | 2.4 | 2.5 | 2.0 |
| SC** | 0.7 | [0.3, 1.5] | 0.7 | [0.4, 1.3] | 0.6 | [0.3, 1.4] | 1.0 | 1.0 | 0.9 |
| SD | 1.1 | [0.5, 2.3] | 1.1 | [0.6, 1.9] | 0.9 | [0.4, 2.0] | 0.8 | 0.8 | 0.7 |
| TN | 0.9 | [0.4, 2.0] | 1.0 | [0.5, 1.7] | 0.8 | [0.3, 1.8] | 0.9 | 1.0 | 0.8 |
| TX | 0.9 | [0.4, 2.0] | 1.0 | [0.5, 1.7] | 0.8 | [0.3, 1.8] | 0.6 | 0.7 | 0.6 |
| UT | 0.5 | [0.3, 1.1] | 0.6 | [0.3, 1.0] | 0.4 | [0.2, 1.0] | 0.7 | 0.7 | 0.6 |
| VA | 1.3 | [0.6, 2.7] | 1.4 | [0.8, 2.3] | 1.1 | [0.5, 2.4] | 0.9 | 1.0 | 0.8 |
| VT | 1.0 | [0.4, 2.1] | 1.0 | [0.6, 1.8] | 0.8 | [0.3, 1.8] | 1.4 | 1.5 | 1.2 |
| WA* | 1.3 | [0.6, 2.7] | 1.4 | [0.8, 2.3] | 1.1 | [0.5, 2.4] | 1.4 | 1.4 | 1.1 |
| WI | 0.7 | [0.3, 1.4] | 0.7 | [0.4, 1.2] | 0.6 | [0.2, 1.3] | 0.9 | 0.9 | 0.7 |
| WV | 0.7 | [0.3, 1.6] | 0.7 | [0.4, 1.3] | 0.6 | [0.2, 1.4] | 0.4 | 0.4 | 0.4 |
| WY | 0.3 | [0.1, 0.6] | 0.3 | [0.2, 0.5] | 0.2 | [0.1, 0.6] | 0.7 | 0.7 | 0.6 |

Notes: Columns (1) to (6) report the estimates and heteroskedasticity robust 95% confidence intervals for population prevalence of COVID-19 on April 7 based on the methodology described in Section 2. Columns (7) to (9) report the the average estimates for population prevalence of COVID-19 from March 31 to April 7. Columns (3), (4) and (8) report results based on models that include state fixed effects. Columns (5), (6), and (9) report results based on models that restrict the sample to observations for which the share of positive cases was less than 0.5. In cases of incomplete testing data on April 7, population prevalence is reported for the closest day: \* indicates prevalence on April 6, \*\* indicates prevalence on April 5, and \*\*\* indicates prevalence on March 31.

Table 2, col. (4) reports the average estimated population prevalence for the period March 31 to April 7. These averages mitigate sampling error in the daily prevalence estimates, which depend on the observed share of positive tests on any particular day. The average estimates are similar to the April 7 estimates, albeit generally smaller in magnitude, suggesting continued spread of the disease in many states.

In Table 3, we examined the robustness of the main estimates. To begin, we estimated modified versions of equation (4) that include state fixed effects. These models allow for an exponential trend in infection rates, thereby addressing concerns that underlying disease prevalence may evolve from one day to the next. We allowed each state to have its own specific intercept to capture the fact that the trends may differ depending on the local conditions. The results (reported in cols. 2 and 7) are virtually identical to the baseline estimates. Moreover, the augmented model tends to produce more precise confidence intervals.

**Table 3:** Diagnosed Cases and Estimated Total Cases of COVID-19

| State | Positive<br>COVID-19 Tests,<br>by April 12 | Estimated Total<br>COVID-19 Cases | Ratio of Total Cases<br>to Positive Tests<br>(2)/(1) | COVID-19 Tests<br>per 1,000<br>Population |
| --- | --- | --- | --- | --- |
|  | (1) | (2) | (3) | (4) |
| AK | 272 | 3,177 | 11.7 | 11.0 |
| AL | 3,525 | 44,155 | 12.5 | 4.4 |
| AR | 1,280 | 16,460 | 12.9 | 6.5 |
| AZ | 3,539 | 45,434 | 12.8 | 5.8 |
| CA | 21,794 | 353,000 | 16.2 | 4.8 |
| CO | 6,893 | 106,505 | 15.5 | 6.1 |
| CT | 12,035 | 148,252 | 12.3 | 11.6 |
| DC | 1,875 | 20,843 | 11.1 | 15.1 |
| DE | 1,479 | 14,779 | 10.0 | 11.4 |
| FL | 19,355 | 274,117 | 14.2 | 8.5 |
| GA | 12,452 | 215,306 | 17.3 | 5.1 |
| HI | 486 | 6,179 | 12.7 | 12.7 |
| IA | 1,587 | 22,678 | 14.3 | 5.6 |
| ID | 1,407 | 27,105 | 19.3 | 8.0 |
| IL | 20,852 | 287,087 | 13.8 | 7.9 |
| IN | 7,928 | 128,568 | 16.2 | 6.3 |
| KS | 1,337 | 19,110 | 14.3 | 4.5 |
| KY | 1,840 | 18,328 | 10.0 | 5.5 |
| LA | 20,595 | 310,465 | 15.1 | 22.4 |
| MA | 25,475 | 236,752 | 9.3 | 16.8 |
| MD | 8,225 | 102,114 | 12.4 | 8.2 |
| ME | 633 | 7,165 | 11.3 | 5.0 |
| MI | 24,638 | 441,486 | 17.9 | 8.0 |
| MN | 1,621 | 18,007 | 11.1 | 6.6 |
| MO | 4,160 | 69,549 | 16.7 | 7.4 |
| MS | 2,781 | 32,330 | 11.6 | 7.2 |
| MT | 387 | 5,138 | 13.3 | 8.3 |
| NC | 4,520 | 66,830 | 14.8 | 5.9 |
| ND | 308 | 3,507 | 11.4 | 13.6 |
| NE | 791 | 10,821 | 13.7 | 5.5 |
| NH | 929 | 16,792 | 18.1 | 8.0 |
| NJ | 61,850 | 672,314 | 10.9 | 14.3 |
| NM | 1,174 | 13,696 | 11.7 | 13.7 |
| NV | 2,836 | 36,864 | 13.0 | 8.0 |
| NY | 188,694 | 1,644,119 | 8.7 | 23.7 |
| OH | 6,604 | 100,221 | 15.2 | 5.4 |
| OK | 1,970 | 38,186 | 19.4 | 5.8 |
| OR | 1,527 | 21,994 | 14.4 | 7.1 |
| PA | 22,833 | 302,535 | 13.2 | 9.8 |
| RI | 2,665 | 25,081 | 9.4 | 19.2 |
| SC | 3,319 | 51,737 | 15.6 | 6.1 |
| SD | 730 | 7,205 | 9.9 | 9.7 |
| TN | 5,308 | 64,366 | 12.1 | 10.3 |
| TX | 13,484 | 187,963 | 13.9 | 4.3 |
| UT | 2,303 | 22,403 | 9.7 | 13.8 |
| VA | 5,274 | 80,574 | 15.3 | 4.7 |
| VT | 727 | 8,867 | 12.2 | 15.8 |
| WA | 10,224 | 103,188 | 10.1 | 12.3 |
| WI | 3,341 | 50,662 | 15.2 | 6.7 |
| WV | 611 | 7,724 | 12.6 | 9.1 |
| WY | 261 | 3,921 | 15.0 | 9.4 |

*Notes:* Columns (1) reports the cumulative number of positive COVID-19 tests by April 12. Column (2) reports the total number of COVID-19 cases implied by the average estimated population prevalence from March 31 to April 7 (Table 2, col. 4). Column (4) reports the cumulative number of COVID-19 tests by April 12 per 1,000 population.

We explored the sensitivity of the results to excluding days in which a large fraction of tests were positive. This specification addresses concerns that the functional form of the estimating equation may differ in settings in which the share of positive was large, due to the approximation in equation (2). We restricted the sample to observations in which fewer than 50% of tests were positive, and re-estimated equation (4). Table 3, cols. 5,6,9 report the results. Although the sample size is reduced, the predicted infection rates are similar in magnitude to the baseline estimates and have similar confidence intervals.

In Table 4, we explored the relationship between the number of diagnosed cases and total population COVID-19 infections implied by our estimation procedure. We compared the average population infection rates from March 31 to April 7 to the total number of diagnosed cases by April 12. Because many individuals may not seek testing until the onset of symptoms, the latter date was chosen to capture the virus’s typical five day incubation period [19, 20]. Column (1) reports the total diagnosed cases by April 12; column (2) reports the total number of COVID-19 cases implied by the estimates reported in Table 2 (col. 4); and column (3) presents the ratio of total cases to diagnosed cases.

The results reveal widespread undetected population infection. Nationwide, we found that for every identified case there were 12 total infections in the population. There were significant cross-state differences in these ratios. In New York, where more than two percent of the population had been tested, the ratio of total cases to positive diagnoses was 8.7, the lowest in the nation. Meanwhile, Oklahoma had the highest ratio in the country (19.4), and tested less than 0.6 percent of its population.

Figure 2 presents a bivariate scatter plot between the ratio of total COVID-19 cases per diagnosis and cumulative per capita testing by April 12. The negative relationship (corr = -0.51) indicates that relative differences in state testing do not simply reflect a response to geographic differences in pandemic severity. Instead, the patterns suggest that states that expanded testing capacity more broadly were better able to track population prevalence.

**Figure 2:**
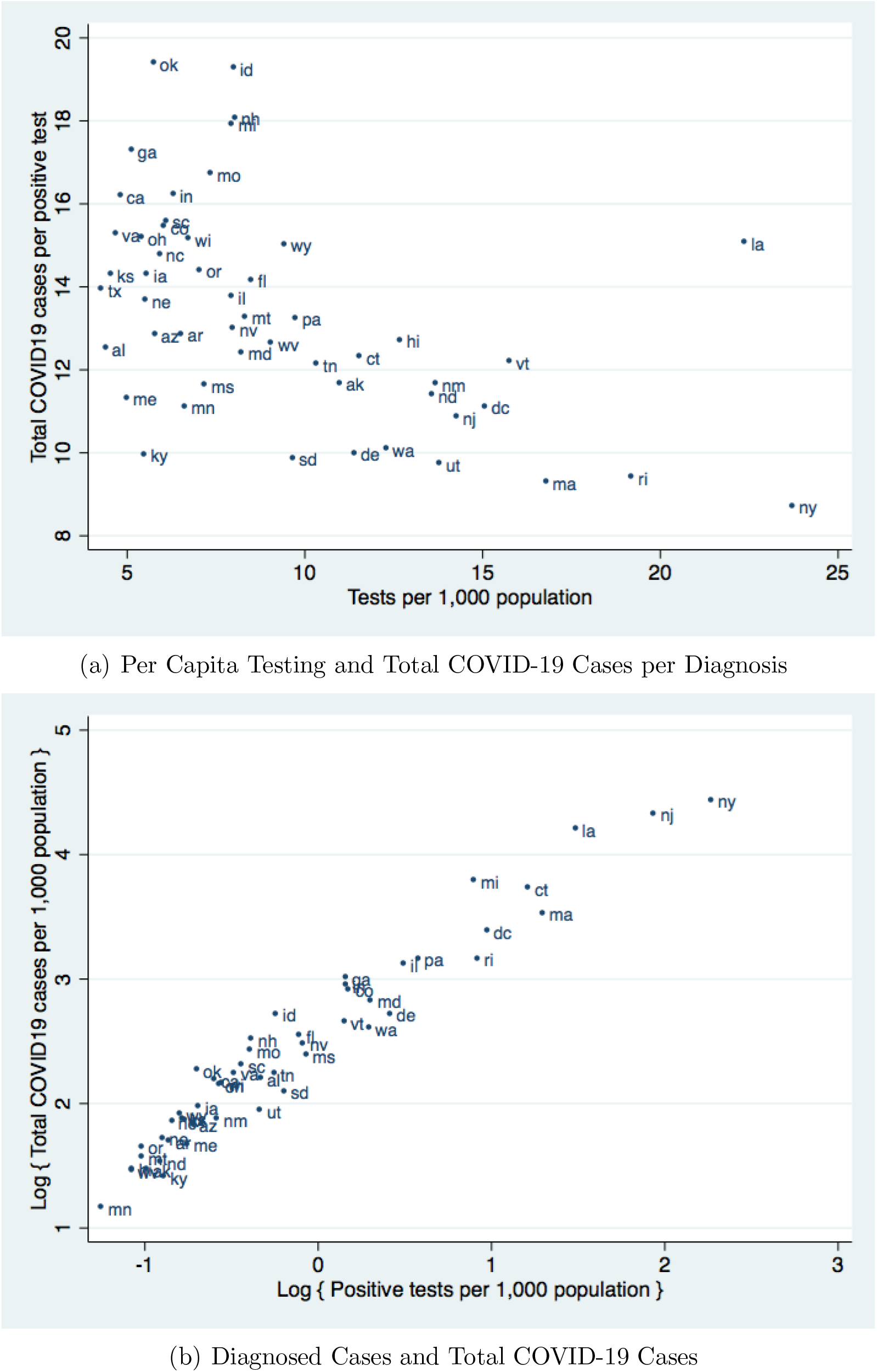
Testing and Population COVID-19 Infection Rates across States. *Notes:* (a) This figure presents the bivariate relationship between per capita testing and the ratio of total COVID-19 cases per diagnosis. Tests per 1,000 population are based on the cumulative number of tests by April 12. The ratio is the total number of COVID-19 cases, derived from the average estimated population prevalence from March 31 to April 7, divided by the cumulative number of positive tests by April 12. (b) This figure presents the bivariate relationship between log positive tests per capita and log total COVID-19 cases per capita. Positive tests per 1,000 population are based on the cumulative number of positive tests by April 12. The total number of COVID-19 cases is derived from the average estimated population prevalence from March 31 to April 7.

Figure 3 documents a positive relationship between per capita COVID-19 diagnoses and population prevalence. The similarity between these two series is notable, given that our estimates were derived from an entirely different source of variation from the cumulative case counts. Nevertheless, observed case counts do not perfectly predict overall population prevalence. For example, despite similar rates of reported positive tests, Michigan had roughly twice as many per capita infections as Rhode Island. These differences can partly be explained by the fact that nearly two percent of the population in Rhode Island had been tested by April 12, whereas fewer than one percent had been tested in Michigan. Together, these findings suggest that differences in state-level policies towards COVID-19 testing may mask important differences in underlying disease prevalence.

## 4 Discussion

The high proportion of asymptomatic and mild cases coupled with limitations in laboratory testing capacity has created large uncertainty regarding the extent of the COVID-19 outbreak among the general population. As a result, key elements of virus’ clinical and epidemiological characteristics remain poorly understood. This uncertainty has also created significant challenges to policymakers who must trade off the potential benefits from non-pharmaceutical interventions aimed at curbing local transmission against their substantial economic and social costs.

A number of recent studies have sought to estimate COVID-19 disease prevalence and mortality in the United States and internationally [21–26]. One approach has been based on variants of the Susceptible Infectious Removed (SIR) model, in which parameters are “calibrated” to the specific characteristics of the SARS-CoV-2 pandemic to estimate current and future infections. A challenge for this approach is the large uncertainty regarding the relevant parameter values for the virus, and the fact that the parameter values will evolve as societies take different measures to reduce transmission. Other research has relied on Bayesian modelling to infer past disease prevalence from observed COVID-19 deaths, and apply SIR models to forecast current infection rates. This approach requires fewer assumptions regarding the underlying parameter values. Nevertheless, because these models ‘scale up’ observed deaths to estimate population infections, small differences in the assumed case fatality will have substantial effects on the results. This poses a challenge for estimation, given that there is considerable uncertainty regarding the case fatality rate, which may vary widely across regions due to local demographics and environmental conditions [27–31]. Moreover, to the extent that there is significant undercounting in the number of COVID-19 related deaths [32, 33], these estimates may fail to capture the full extent of population infection.

In this paper, we developed a new methodology to estimate population disease prevalence when testing is non-random. Our approach builds on a standard econometric technique that have been used to address sample selection bias in a variety of different settings. Our estimation strategy offers several advantages over existing methods. First, the analysis has minimal data requirements. The three variables used for estimation – daily infections, daily number of tests, and total population – are widely reported across a large number of countries and subnational districts. Second, the model identification is transparent and depends only on a simple exclusion restriction assumption that daily changes in the number of conducted tests must be uncorrelated with underlying changes in population disease prevalence. This assumption is likely to hold in many jurisdictions where constraints on capacity are a primary determinant of testing.

We used this framework to estimate disease prevalence across U.S. states. We estimated substantial population infection that exceeded the observed rates of positive tests by factors of 8 to 19. These results are consistent with recent evidence suggesting that there may be widespread undetected infection across many regions of the U.S. [26]. Our findings are comparable to previous studies on U.S. population prevalence that find ratios of population infection to positive tests ranging from 5 to 10 by mid-March [22, 25]. Despite a dramatic expansions in testing capacity in the intervening weeks, the vast majority of COVID-19 cases remain undetected.

Our results are comparable to recent estimates of population prevalence in a number of European countries [21]. We found a nationwide 1.9 percent infection rate in early April, which is similar to the estimated prevalence in Austria (1.1%), Denmark (1.1%), and the United Kingdom (2.7%) as of March 28. Meanwhile, Germany’s 0.7% infection rate would rank in the lowest tercile of prevalence among U.S. states. The highest rates of infection in New York (8.5%), New Jersey (7.6%), and Louisiana (6.7%) are still lower than the estimated rates in Italy (9.8%) and Spain (15%). Given the rapidly expanding availability of high frequency testing data at both the national and subnational level, in future research we plan to apply this methodology to compare infection rates across a broader spectrum of countries.

There are several limitations to our study, which should be taken into account when interpreting the main findings. First, the estimation results depend on several functional form assumptions including a constant exponential growth rate in new infections and the specific functions governing how the number of available tests affect individual testing probability. As more data on testing become available, the increased sample sizes will allow future studies to impose weaker functional form assumptions through either semi- or non-parametric approaches. Second, our analysis required an assumption that the underlying sample selection process was similar across observations. To the extent that decisions regarding *who* to test, conditional on the number of available tests, diverged across states or changed within states over the sample period, our model may be misspecified. Finally, our analysis depends on the quality of diagnostic testing, and systematic false negative test results may affect the population disease prevalence estimates [34–36].^4^

As countries continue to struggle against the ongoing coronavirus pandemic, informed policymaking will depend crucially on timely information on infection rates across different regions. Randomized population-based testing can provide this information, however, given the constraints on supplies, this approach has largely been eschewed in favor of targeted testing towards high risk groups. In this paper, we developed a new approach to estimate population disease prevalence when testing is non-random. The estimation procedure is straightforward, has few data requirements, and can be used to estimate disease prevalence at various jurisdictional levels.

## Data Availability

Data is publicly available (see manuscript for details).

## Contributions

DB, RG, and JL conceptualized the study, analyzed the data, and drafted and finalized the manuscript. All authors approved of the final version of the manuscript.

## Acknowledgements

This study was supported by funding from the Social Sciences and Humanities Research Council (Grant: SSHRC 430-2017-00307).

## A Appendix: Tables and Figures

**Table A.1:**
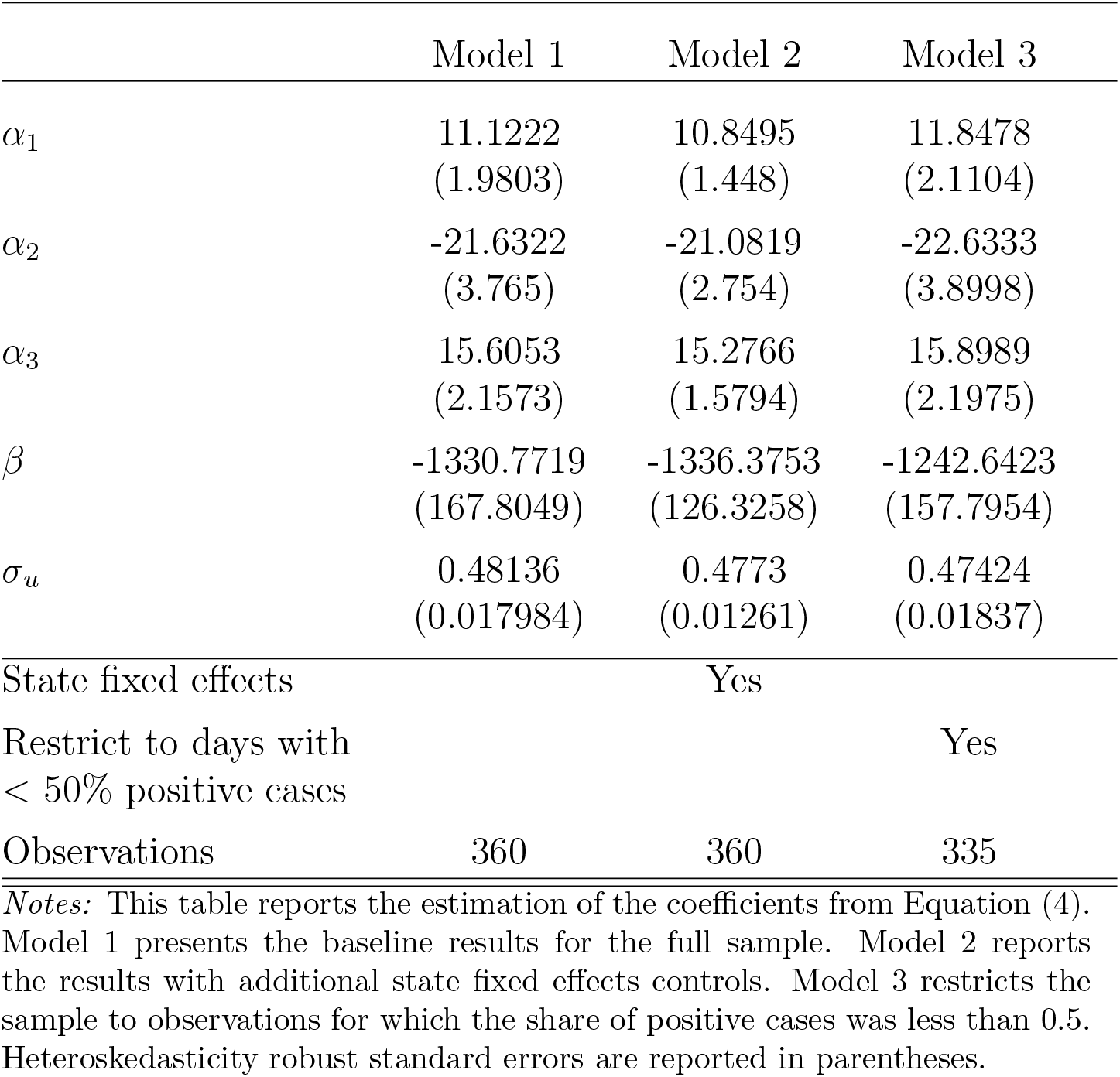
Coefficient Estimates from Equation (4)

## Footnotes

1 Notable exceptions include the universal testing of passengers on the Diamond Princess cruise ship, and an ongoing population-based test project in Iceland.

2 The median ratio of negative to positive tests is 7.3 to 1. In the empirical analysis, we assess the sensitivity of the results to this approximation.

3 To the extent that day-to-day changes in testing responded endogenously to changes in disease prevalence, we might actually expect this relationship to be positive. In this scenario, our estimates should be interpreted as a lower bound for sample selection bias.

4 Provided that the rates of misdiagnosis were unrelated to the number of tests, these errors will not bias the coefficient estimates, but may reduce precision through classical measurement error [37].

